# Study protocol for preoperative classification using integrated screening and short-course neoadjuvant BRAF/MEK inhibition in newly diagnosed papillary craniopharyngioma (the PRECISE-PCP study): a prospective single-arm study

**DOI:** 10.64898/2026.05.08.26351826

**Authors:** Zhen Ye, Guoqing Wu, Haoqin Jiang, Xufang Gu, Ruofan Huang, Yuting Wang, Zengyi Ma, Zhao Ye, Nidan Qiao, Yue Wu, Weiwei Wang, Haixia Cheng, Hong Chen, Hongying Ye, Yongfei Wang, Zhaoyun Zhang, Ming Guan, Yao Zhao, Qilin Zhang

## Abstract

**Introduction:** Craniopharyngioma (CP) comprises two distinct histological subtypes, adamantinomatous craniopharyngioma (ACP) and papillary craniopharyngioma (PCP), which are often challenging to distinguish preoperatively. Approximately 95% of PCP harbor the BRAF V600E mutation, whereas ACP lacks this alteration, making PCP uniquely sensitive to BRAF and MEK inhibition. However, in the absence of a reliable preoperative classification strategy, targeted therapy has been limited to recurrent disease or to cases with histological confirmation. This study aims to describe and prospectively evaluate a pragmatic preoperative classification strategy and short-course neoadjuvant BRAF and MEK inhibition followed by surgery in newly diagnosed, preoperatively classified PCP.

**Methods and analysis:** This is a prospective, single-arm, open-label study. Patients with newly diagnosed craniopharyngioma will be screened using an integrated preoperative strategy combining imaging-based prediction and selective cerebrospinal fluid (CSF) cell-free DNA testing for BRAF V600E in indeterminate cases. Twelve participants preoperatively predicted as PCP and BRAF V600E positive will receive dabrafenib 150 mg twice daily plus trametinib 2 mg once daily for up to three 28-day cycles, followed by transnasal endoscopic surgery. Assessments are scheduled at days 7, 14, 28, 56, and 84 until surgery. The primary endpoint is objective response rate, assessed by contrast-enhanced MRI using RANO 2.0 criteria. Secondary outcomes include progression-free survival, local disease control, endocrine outcomes of the hypothalamic-pituitary-adrenal and hypothalamic-pituitary-thyroid axes, visual and cognitive outcomes, postoperative diabetes insipidus, surgical complexity, and concordance between the preoperative classification strategy and postoperative pathology and BRAF V600E status. Exploratory analyses will evaluate treatment-related changes in tumor vascularity, tissue characteristics, and post-treatment molecular alterations in tumor tissue.

**Ethics and dissemination:** This protocol has been approved by the Ethics Committee of Huashan Hospital, Fudan University (KY2024-028). Written informed consent will be obtained from all participants. Results will be disseminated through peer-reviewed publications and scientific conferences.

**Trial registration number:** ChiCTR2400081636

**STRENGTHS AND LIMITATIONS OF THIS STUDY:** *Strength:*

➢ This study proposes an integrated, clinically applicable preoperative strategy that combines imaging-based prediction with selective cerebrospinal fluid cell-free DNA analysis to identify papillary craniopharyngioma (PCP) prior to surgery.
➢ It prospectively evaluates short-course neoadjuvant BRAF and MEK inhibition in newly diagnosed PCP, addressing a clinically relevant gap in current management.
➢ Standardized, multidimensional assessments are performed across the neoadjuvant, perioperative, and early postoperative periods, capturing radiographic, surgical, endocrine, visual, and cognitive outcomes.

*Limitation:*

➢ The single-arm, open-label design without a surgical control group limits direct comparison with upfront surgery.
➢ Despite the integrated prediction strategy, preoperative misclassification cannot be excluded entirely.

## INTRODUCTION

Craniopharyngiomas are rare epithelial tumors of the sellar and suprasellar region, accounting for approximately 1–4% of intracranial neoplasms. They are classified into adamantinomatous (ACP) and papillary (PCP) subtypes, with PCP comprising about 30–40% of cases. Surgical resection remains the standard treatment. However, the close anatomical relationship of these tumors to the hypothalamus, optic apparatus, and pituitary stalk makes surgery technically demanding and frequently associated with substantial long-term morbidity, including permanent hypopituitarism and neurocognitive impairment^1,2^. Against this background, PCP differs from ACP in that most tumors harbor the BRAF V600E mutation, providing a clear molecular target and the potential to modify the traditional surgery-centered management strategy.

Clinical evidence accumulated to date indicates that BRAF and MEK inhibitors can induce rapid and substantial tumor regression in PCP, with over 90% of patients reaching a 50% reduction of tumor within 3 months^3–6^. Most published experience, however, has focused on patients with recurrent disease or those who have already undergone surgery or radiotherapy^7^. Emerging retrospective data suggest that when BRAF and MEK inhibitors are used earlier in the disease course, particularly in newly diagnosed PCP, tumor shrinkage tends to occur more quickly and to a greater extent than when treatment is initiated after prior interventions^6,8^. These observations suggest that the initial presentation of PCP may represent an optimal therapeutic window in which early targeted intervention could maximize treatment response. Consequently, accurate preoperative identification of PCP is required to enable such an approach.

Several groups have explored imaging-based artificial intelligence approaches to distinguish PCP from ACP. Still, no publicly available model has been widely adopted in clinical practice, and prospective multicenter validation remains limited^9–11^. To address this gap, an imaging-based prediction model has been developed and evaluated using independent cohorts to support preoperative subtype prediction. Because imaging-based prediction cannot achieve perfect accuracy, a subset of patients inevitably falls into a diagnostic grey zone. In such cases, additional non-invasive or minimally invasive approaches are required to improve preoperative confidence. Prior studies have shown that BRAF-mutant intracranial tumors can shed tumor-derived cell-free DNA (cf-DNA) into the cerebrospinal fluid (CSF), allowing detection of BRAF V600E mutations without tissue biopsy^4,12,13^. Accordingly, CSF cf-DNA analysis can serve as a practical adjunct to imaging-based prediction in patients with indeterminate imaging results, enabling a structured preoperative strategy that prioritizes imaging assessment, incorporates molecular confirmation when needed, and avoids routine biopsy.

The availability of preoperative prediction creates the opportunity to evaluate how such information should guide initial treatment decisions prospectively. Key unanswered questions include the objective response rate (ORR) achieved with short-course neoadjuvant BRAF and MEK inhibition in patients with newly diagnosed, preoperatively predicted PCP, as well as the ability of the prediction strategy to identify patients most likely to benefit from this approach. In addition, the optimal management strategy following treatment-induced tumor regression remains unclear. Long-term targeted therapy is often impractical because of economic burden and patient tolerance, raising the question of whether surgical resection after short-course treatment could serve as a definitive therapeutic option. It also remains unknown how short-course neoadjuvant therapy influences surgical complexity and postoperative endocrine outcomes.

These unresolved issues support the need for a prospective, single-arm study to assess the safety, feasibility, and perioperative impact of short-course neoadjuvant BRAF and MEK inhibition in newly diagnosed PCP.

## METHODS and ANALYSIS

This protocol describes a prospective, single-arm, open-label study in patients with newly diagnosed craniopharyngioma who are preoperatively classified as PCP and BRAF V600E–positive using an integrated strategy combining imaging-based prediction and CSF cf-DNA testing. Eligible participants will receive short-course neoadjuvant dabrafenib plus trametinib, with radiographic response assessed before planned surgical management.

### Patient and public involvement

Patients and the public were not involved in the design, conduct, reporting, or dissemination plans of this research.

#### Inclusion criteria

➢ 18-74 years of age.
➢ KPS score ≥70 within 2 weeks before treatment.
➢ Clinically diagnosed craniopharyngioma based on imaging and multidisciplinary assessment.
➢ Preoperatively classified as PCP with BRAF V600E, based on the integrated preoperative screening strategy combining imaging-based prediction and CSF cf-DNA analysis, where applicable.
➢ Lesions were assessed as measurable according to the RANO (Response Assessment in Neuro-Oncology) 2.0 criteria.
➢ Life expectancy ≥ 6 months.
➢ Ability to understand and the willingness to sign a written informed consent document.

#### Exclusion criteria

➢ Simultaneously suffering from other malignant tumors, except for those that have been cured or are in a stable condition.
➢ Pregnant or breastfeeding women.
➢ Participated in other clinical trials within the past six months.
➢ Hemoglobin < 90 g/L, Absolute neutrophil count < 1.5×10^9^/L, platelet count < 80×10^9^/L.
➢ Total Bilirubin > 1.5×ULN, ALT/AST > 2.5×ULN, Ccr < 50 mL/min (determined by Cockcroft-Gault).
➢ Clinically significant cardiovascular disease, including uncontrolled hypertension, recent myocardial infarction, unstable angina, significant arrhythmia, or NYHA class ≥ II heart failure.
➢ Patients who have experienced arterial/venous thrombotic events within the year before screening, such as cerebrovascular accidents (including transient ischemic attacks), deep vein thrombosis (excluding venous thrombosis caused by venous catheterization for prior chemotherapy and judged by the investigator to have fully recovered), and pulmonary embolism, etc.
➢ Patients who have experienced any bleeding event (≥CTCAE 4.0 grade 3) within 4 weeks before screening; those with significant coagulation abnormalities or a tendency to bleed; patients undergoing treatment with warfarin, heparin, or their analogs; the use of low-dose warfarin (1 mg orally, once daily) or low-dose aspirin (daily dose not exceeding 100 mg) for preventive purposes is allowed, provided the international normalized ratio (INR) of prothrombin time is ≤ 1.5.
➢ Patients with multiple factors affecting oral medication (e.g., inability to swallow, chronic diarrhea, intestinal obstruction, etc.), or those requiring treatment with strong CYP3A4 inducers or CYP3A4 inhibitors.
➢ History of uveitis or iritis within the past 4 weeks; history of retinal diseases (such as neurosensory retinal detachment, retinal vein occlusion, or neovascular macular degeneration) within the past 12 months.
➢ History of organ transplantation; history of immunodeficiency or other acquired or congenital immunodeficiency diseases; long-term unhealed wounds or incomplete fracture healing; known chronic lung diseases; history of psychotropic drug abuse with inability to abstain or presence of mental disorders.
➢ Patients were deemed unsuitable for participation in the trial based on the investigator’s judgment.

#### Withdrawal criteria

➢ Subjects who are unwilling or unable to continue participating in the trial.
➢ Incorrectly enrolled subjects, i.e., the subject does not meet the required inclusion/exclusion criteria for the study.
➢ If an intolerable adverse event (AE) or serious adverse event (SAE) occurs, and after dose adjustment, the investigator judges that the risks to the subject of continuing participation outweigh the potential benefits.
➢ Subject is lost to follow-up.

### Screening

#### Imaging-based prediction model

Preoperative MR and CT images were obtained as part of routine clinical evaluation and retrieved from the institutional PACS in DICOM format. CT images were used for visual assessment of intratumoral calcification by experienced neuroradiologists. MRI-based modeling was restricted to conventional T1-weighted and contrast-enhanced T1-weighted sequences to ensure consistency across centers.

After standardized image preprocessing, regions of interest encompassing the sellar and parasellar tumor regions were defined for feature extraction. An imaging-based prediction model was developed using a large single-center cohort for model training, followed by validation in an independent cohort from the same center. The model was further evaluated in a multicenter cohort to assess generalizability. Model inputs included features derived from T1-weighted and contrast-enhanced T1-weighted MRI, combined with manually assessed calcification status and basic clinical variables, including age and sex, to distinguish PCP from ACP. The model generates a continuous imaging-based prediction score (0–1), with higher values indicating a greater likelihood of PCP. Model performance was assessed using standard metrics for discrimination and calibration and subsequently applied in a prospective setting to support preoperative subtype prediction.

#### Lumbar puncture and CSF sample collection

Lumbar puncture for CSF sampling was performed selectively based on imaging-based prediction results. In patients with indeterminate imaging-based prediction scores, CSF analysis was used to improve preoperative confidence. Approximately 3 mL of CSF was collected and processed for cf-DNA extraction according to standard protocols. Detection of the BRAF V600E mutation in CSF cf-DNA was performed using droplet digital PCR (ddPCR). CSF cf-DNA results were integrated with the imaging-based prediction score to support preoperative subtype classification and treatment decision-making.

#### Integration of CSF and imaging model prediction results

Imaging-based prediction served as the primary method for preoperative treatment stratification. Patients with high-confidence imaging-based predictions proceeded directly to treatment decision-making. In cases with indeterminate imaging-based scores, CSF cf-DNA analysis was selectively performed to assess the presence of the BRAF V600E mutation. Imaging-based prediction scores and CSF cf-DNA results were jointly interpreted to determine eligibility for study treatment (Figure 1).

**Figure 1.**
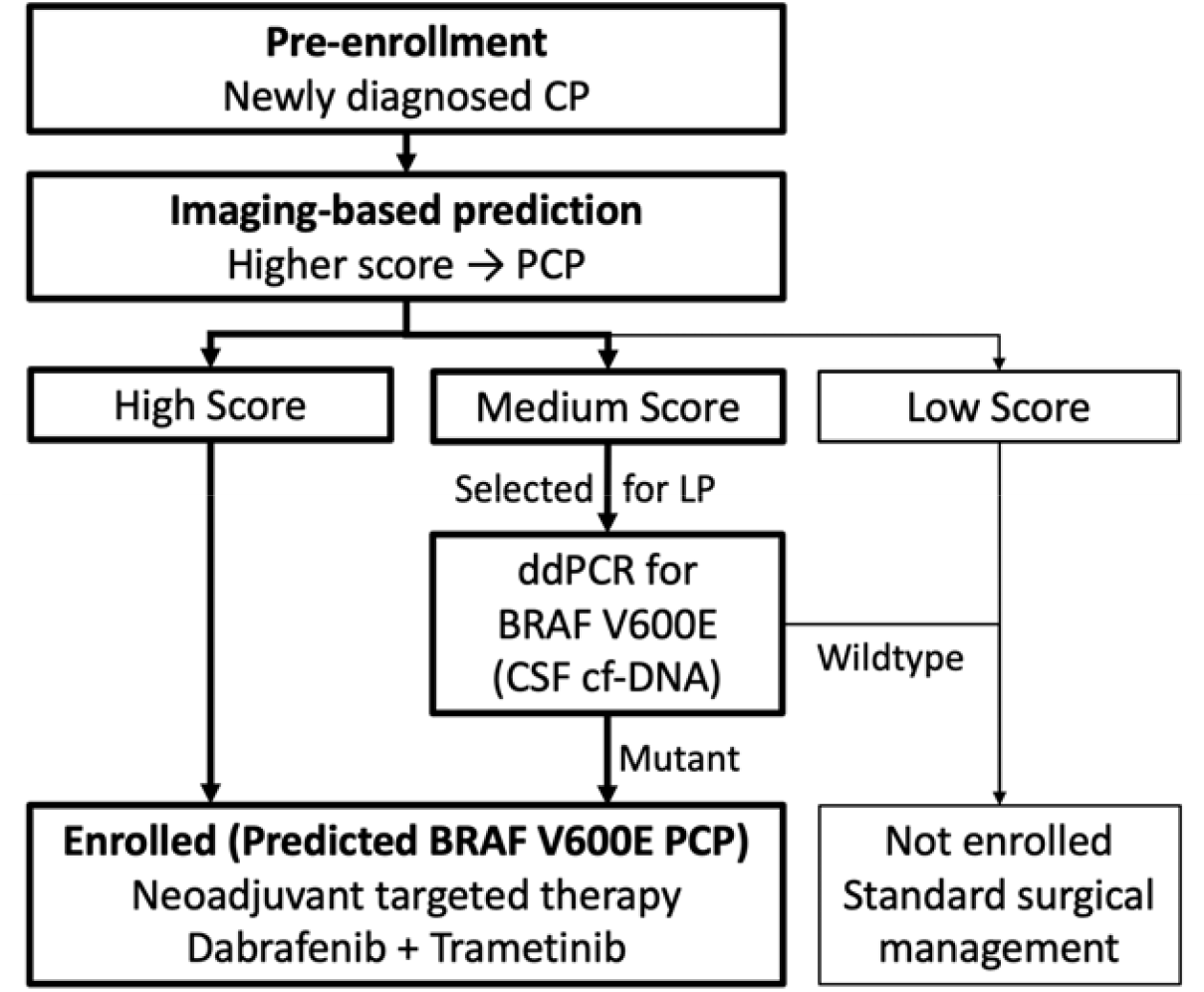
Preoperative screening and enrollment workflow. Workflow illustrating the preoperative screening and enrollment strategy for study treatment (neoadjuvant dabrafenib plus trametinib). Patients with newly diagnosed craniopharyngioma (CP) undergo integrated imaging-based prediction, generating a score in which higher values indicate a greater likelihood of papillary craniopharyngioma (PCP). Patients with high scores were classified as predicted PCP and considered eligible for neoadjuvant therapy. Patients with intermediate scores are selectively referred for lumbar puncture (LP), with droplet digital PCR (ddPCR) analysis of BRAF V600E in cerebrospinal fluid (CSF) cf-DNA used to refine subtype classification. Patients classified as predicted PCP are enrolled to receive short-course neoadjuvant dabrafenib plus trametinib, whereas patients with low scores or BRAF wild-type results proceed with standard surgical management. CP, craniopharyngioma; PCP, papillary craniopharyngioma; LP, lumbar puncture; CSF, cerebrospinal fluid; cf-DNA, cell-free DNA; ddPCR, droplet digital PCR.

### Intervention

Eligible patients with preoperatively predicted PCP will receive neoadjuvant dabrafenib 150 mg orally twice daily in combination with trametinib 2 mg orally once daily. Each treatment cycle consists of 28 days. Treatment will be administered for up to three cycles, with treatment duration determined by radiographic response and subsequent surgical decision-making. Targeted therapy will be discontinued at the time of surgical intervention (Figure 2).

**Figure 2.**
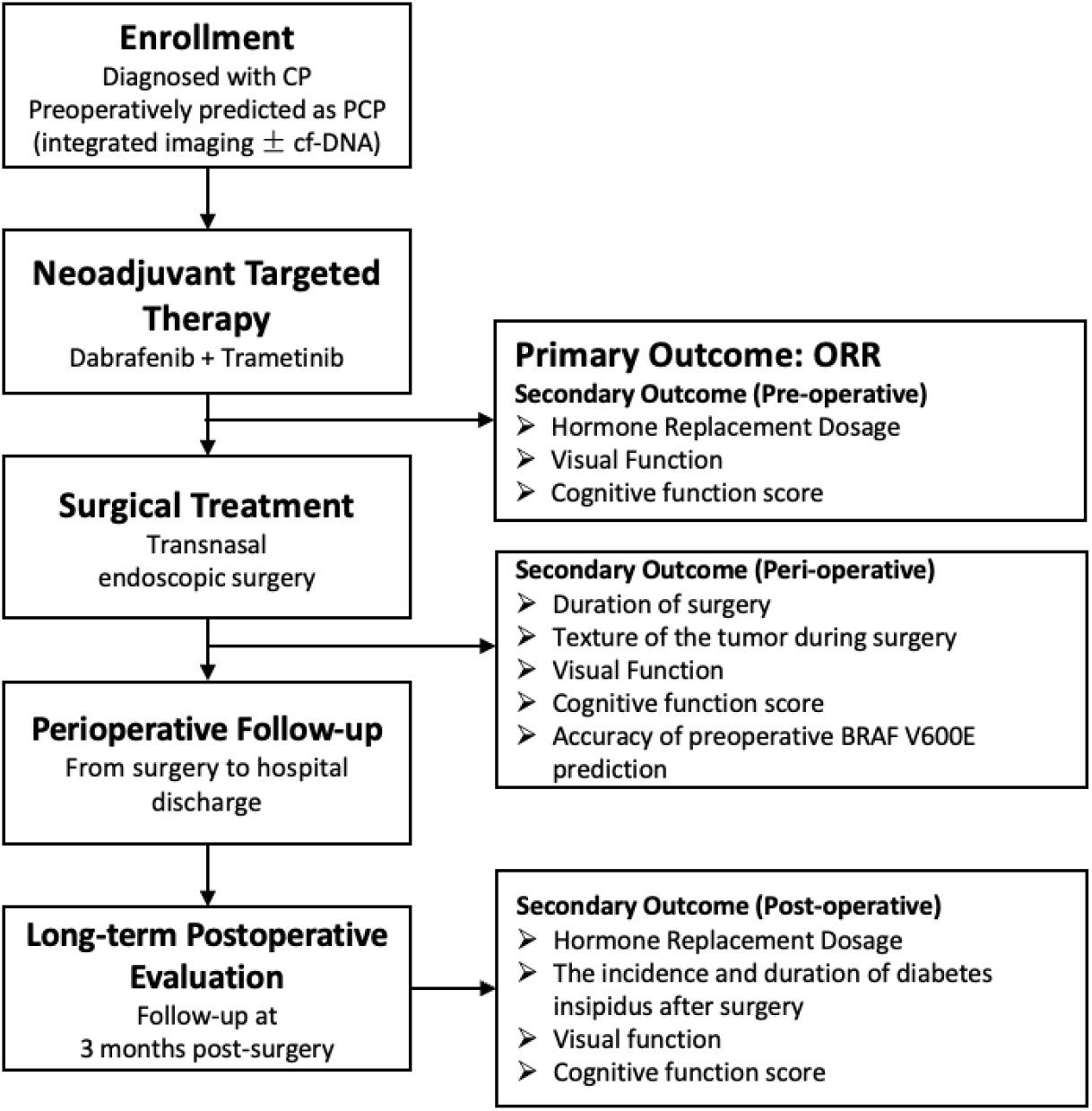
Study workflow and outcome assessment timeline. Study flow illustrating enrollment, treatment, surgery, and follow-up. Patients diagnosed with craniopharyngioma (CP) are preoperatively classified as papillary craniopharyngioma (PCP). Eligible patients receive short-course neoadjuvant dabrafenib plus trametinib, followed by transnasal endoscopic surgery. The primary outcome is objective response rate (ORR). Secondary outcomes are assessed at predefined perioperative and postoperative time points, including surgical, endocrine, visual, cognitive, and predictive accuracy measures. CP, craniopharyngioma; PCP, papillary craniopharyngioma; CSF, cerebrospinal fluid; cf-DNA, cell-free DNA; ORR, objective response rate.

### Radiologic Assessment

This single-center study used independent radiologic review for all imaging assessments. Two board-certified radiologists with expertise in skull base tumors independently evaluated all imaging studies and were blinded to clinical data and treatment outcomes. In cases of discordant assessments regarding tumor volume change or response classification, a third radiologist performed adjudication and provided the final determination. All imaging evaluations were conducted according to prespecified, standardized criteria defined in the study protocol to ensure consistency and reliability. All radiologic assessments were performed according to a predefined analysis plan and were not influenced by clinical decision-making.

### Follow-up

Follow-up assessments were scheduled at days 7, 14, 28, 56, and 84 after initiation of neoadjuvant therapy and continued until surgical intervention, at which point targeted therapy was discontinued. For patients who underwent surgery, an additional follow-up visit was conducted 3 months postoperatively.

Radiographic response was evaluated at each follow-up using contrast-enhanced MRI and compared with baseline imaging, consistent with RANO-based assessment^14^. If MRI demonstrated significant tumor change, defined as a tumor size decrease of ≥50% or an increase of ≥ 25%, surgical intervention was initiated immediately. In patients without significant radiographic change, defined as a tumor size decrease of <50% and an increase of <25%, one 28-day treatment cycle was completed before proceeding to surgical treatment. For patients who met predefined criteria for surgical intervention but declined surgery, continued follow-up was conducted with close monitoring. Decisions regarding the continuation or discontinuation of targeted therapy were made on an individual basis based on clinical assessment within the predefined study framework.

## Outcomes

### Primary outcome

The primary outcome is the objective response rate (ORR), defined as the proportion of patients achieving complete response (CR) or partial response (PR) at the prespecified primary response assessment, defined as the contrast-enhanced MRI performed at the end of neoadjuvant therapy (Day 84). For participants who undergo surgery earlier than Day 84, the last MRI assessment prior to surgery will serve as the primary response assessment. Tumor response will be assessed using contrast-enhanced sellar MRI according to RANO 2.0 criteria, based on two-dimensional measurements on the slice/plane showing the largest tumor cross-section. The timing of the primary response assessment was prespecified and independent of treatment response.

### Secondary outcomes

Secondary outcomes include:

➢ **Progression-Free Survival (PFS):** defined as the time from treatment initiation to radiographic progression or death from any cause, whichever occurs first. Given the protocol-mandated surgery, PFS is assessed within the preoperative window; surgery marks the end of observation rather than an event. Therefore, PFS is interpreted as a descriptive endpoint rather than a measure of long-term disease control.
➢ **Local disease control rate:** defined as the proportion of patients achieving CR, PR, or stable disease (SD).
➢ **Endocrine outcomes:** changes in replacement therapy requirements for the hypothalamic–pituitary–adrenal (HPA) and hypothalamic–pituitary–thyroid (HPT) axes before treatment, before surgery, and at 3 months postoperatively.
➢ **Visual outcomes:** changes in visual acuity and visual field function assessed at baseline, before surgery, and 3 months after surgery.
➢ **Cognitive function:** changes in Mini-Mental State Examination (MMSE) scores assessed at baseline, before surgery, and 3 months postoperatively.
➢ **Accuracy of preoperative prediction:** concordance between postoperative tumor pathology and BRAF V600E mutation status and the integrated preoperative prediction strategy.
➢ **Postoperative diabetes insipidus:** incidence and duration, assessed by 24-hour fluid balance, serum sodium levels, and desmopressin requirements.
➢ **Surgical complexity:** operative duration as a surrogate marker of surgical difficulty.

### Exploratory objectives

Exploratory analyses will evaluate changes in tumor vascularity and tissue characteristics following neoadjuvant therapy, as well as post-treatment molecular alterations in tumor tissue, aiming to generate hypotheses for future mechanistic studies.

### Sample size rationale

This study is designed as a prospective, single-arm, exploratory trial evaluating short-course neoadjuvant BRAF and MEK inhibition in patients with newly diagnosed craniopharyngioma who are preoperatively predicted to have PCP with BRAF V600E. The primary endpoint is the objective response rate (ORR).

Based on published data in PCP treated with BRAF and MEK inhibitors, an ORR of approximately 80% has been reported. A Simon two-stage minimax design was therefore adopted to assess whether short-course neoadjuvant therapy demonstrates sufficient antitumor activity to warrant further investigation. The null hypothesis assumes an ORR of ≤50%, which would be considered clinically insufficient, whereas the alternative hypothesis assumes an ORR ≥80%. The type I error was set at 0.05 with a power of 80%.

Using PASS software (version 21), the minimax design requires 6 patients in the first stage; if fewer than 3 patients achieve an objective response, the trial will be terminated for futility. If the study proceeds to the second stage, a total of 12 patients will be enrolled.

Given the low incidence of craniopharyngioma and the proportion of PCP among newly diagnosed cases, approximately 40 patients with newly diagnosed craniopharyngioma are expected to be screened using the integrated preoperative prediction strategy, yielding an estimated 12 patients eligible for neoadjuvant targeted therapy. This sample size satisfies the requirements of the Simon two-stage design and is appropriate for an initial exploratory evaluation.

## Statistical analysis

A detailed statistical analysis plan (SAP) will be developed prior to database lock. All statistical analyses will be conducted using SAS software (version 9.4 or higher).

The primary endpoint, objective response rate (ORR), will be summarized as the proportion of patients achieving complete response (CR) or partial response (PR), with corresponding two-sided 95% confidence intervals calculated using the exact binomial method. The Simon two-stage design will be applied as prespecified to determine whether the observed ORR meets the criteria for study continuation or termination.

Secondary endpoints will be analyzed descriptively. Descriptive analyses may include exploratory within-subject comparisons, without adjustment for multiplicity. Time-to-event outcomes, including progression-free survival (PFS), will be summarized descriptively. Given the short planned preoperative treatment window and the protocol-mandated transition to surgery, the number of progression or death events prior to surgery is expected to be very low; therefore, Kaplan-Meier estimates and median PFS will be reported only if estimable. In such cases, PFS will be reported using event counts and the distribution of observed follow-up time (time at risk) from treatment initiation to progression/death, surgery, or last assessment, as applicable. The date of surgery will be treated as the end of the preoperative observation period rather than a censoring event for survival estimation. Continuous variables will be summarized using descriptive statistics, including mean, standard deviation, median, interquartile range, minimum, and maximum. Categorical variables will be summarized using counts and percentages. Any exploratory statistical tests will be two-sided with an alpha level of 0.05 and interpreted descriptively. Given the exploratory nature of this study, no adjustment for multiple comparisons will be applied to secondary or exploratory analyses.

## DISCUSSION

This study describes a prospective, single-arm clinical trial designed to evaluate a structured preoperative prediction strategy and short-course neoadjuvant targeted therapy in patients with newly diagnosed papillary craniopharyngioma (PCP). By integrating imaging-based prediction with selective cerebrospinal fluid (CSF) cell-free DNA testing for BRAF V600E, the study aims to enable accurate preoperative identification of PCP and to prospectively assess tumor response, surgical outcomes, and early functional effects of neoadjuvant BRAF and MEK inhibition.

The management of papillary craniopharyngioma (PCP) has evolved with the recognition that most tumors harbor the BRAF V600E mutation and respond to BRAF and MEK inhibitors^4,6,12^. Yet, initial treatment decisions for newly diagnosed disease still rely largely on surgery because no standardized preoperative identification strategy exists. A key contribution of this study is the establishment of an integrated preoperative prediction framework tailored to newly diagnosed craniopharyngioma. While imaging-based models can distinguish PCP from adamantinomatous craniopharyngioma, their performance may be limited by inter-institutional variability^10,11^; the selective incorporation of CSF cf-DNA analysis in indeterminate cases improves diagnostic confidence without routine biopsy^13^.

Beyond preoperative identification, this study prospectively evaluates short-course neoadjuvant BRAF and MEK inhibition in newly diagnosed PCP, an area that remains poorly defined. By systematically assessing treatment response, perioperative outcomes, and postoperative endocrine, visual, and cognitive function, this study aims to clarify whether neoadjuvant therapy can reduce tumor burden, facilitate surgery, and support a definitive treatment strategy following short-course targeted therapy.

This study has inherent limitations related to its single-arm, open-label design and limited sample size, which reflect the rarity of PCP. The absence of a control group limits direct comparison with upfront surgery, and misclassification remains possible despite the integrated prediction strategy. Further multicenter studies with larger cohorts will be required to validate these findings and inform future treatment strategies.

In summary, this study describes a structured preoperative classification approach and a prospective single-arm design for evaluating short-course neoadjuvant targeted therapy in newly diagnosed PCP. The findings will provide preliminary evidence to inform future studies and clinical trial design.

## Supporting information

SAP

## Data Availability

All data produced in the present study are available upon reasonable request to the authors.

