## Supplementary material for "Study protocol for preoperative classification using integrated screening and short-course neoadjuvant BRAF/MEK inhibition in newly diagnosed papillary craniopharyngioma (the PRECISE-PCP study): a prospective single-arm study": SAP

### Statistical Analysis Plan

| | |
|---|---|
| **Clinical Trial Protocol Title** | **Preoperative classification using integrated screening and short-course neoadjuvant BRAF/MEK inhibition in newly diagnosed papillary craniopharyngioma (the PRECISE-PCP study): a prospective single-arm study** |
| **Protocol Number** | **KY2024-028** |
| **Investigational Product(s)** | **Dabrafenib tablets (specification: 75mg); Trametinib tablets (specification: 2mg)** |
| **Sponsor** | **Huashan Hospital, Fudan University** |
| **Statistical Analysis Unit** | **Shanghai Zenith Data Technology Co., Ltd** |
| **Zenith Project Number** | **HSYY-HJJB-ⅡT01** |
| **SAP Version Number** | **V1.0** |
| **SAP version date** | **13 March 2026** |

#### Signature Page

**I sign here to confirm that the statistical analysis of the clinical trial data described in the statistical analysis plan (V1.0, 13 March 2026) includ es the statistical analysis content specified in the plan, and I agree with th e statistical analysis methods and output format of the statistical analysis r eport form specified in the statistical analysis plan.**

---

Statistical analysis Unit: Shanghai Zenith Data Technology Co., Ltd

| | | |
|---|---|---|
| Statistician: Meina Wu | Signature: Meina Wu | Date: Mar, 16, 2026 |
| Review Statistician: Rui Song | Signature: Rui Song | Date: Mar, 16, 2026 |

### Signature Page

**I sign here to confirm that the statistical analysis of the clinical trial data described in the statistical analysis plan (V1.0, 13 March 2026) includ es the statistical analysis content specified in the plan, and I agree with th e statistical analysis methods and output format of the statistical analysis r eport form specified in the statistical analysis plan.**

Sponsor: Huashan Hospital, Fudan University

Principal Investigator: Yao Zhao Signature: Yao Zhao Date: Mar. 16, 2026

#### List of Abbreviations

| Abbreviation | Term |
|---|---|
| AE | Adverse Event |
| ATC | Anatomical, therapeutic and chemical classification of drugs |
| CR | Complete response |
| CV | Coefficient of variation |
| ECG | 12-lead electrocardiogram |
| EVS | Evaluable Set |
| FAS | Full Analysis Set |
| GM | Geometric mean |
| Max | Maximum |
| Mean | Arithmetic mean |
| Median | Median |
| MedDRA | Regulatory Activity Medical Dictionary |
| Min | Minimum |
| NMPA | National Medical Products Administration |
| PD | Disease progression |
| PFS | Progression-free survival |
| PR | Partial Response |
| PT | MedDRA Dictionary Preferred Terms |
| QTc | Corrected QT interval |
| Q1 | The first quartile |
| Q3 | The third quartile |
| SAE | Serious Adverse event |
| SD | Standard deviation |
| SD | Stable Disease |
| SOC | MedDRA Dictionary System Organ Classification |
| SS | Safety data set |
| WHOCC | WHO Collaborating Centre for Drug Statistics Methodology |

### 1 Introduction

This is a single-arm, open-label study evaluating experimental drug treatment in patients predicted to have papillary craniopharyngioma. To evaluate the effectiveness of preoperative integrated predictive models, the feasibility and necessity of preoperative use of targeted drugs, and the timing of surgery after medication in patients with new-onset craniopharyngioma. This statistical analysis plan will describe the statistical analysis methods and data processing principles of this trial, and analyze and report the efficacy and safety data of this trial.

The statistical analysis plan is based on the clinical study protocol on the efficacy and safety of preoperative targeted adjuvant therapy in patients with new-onset papillary craniopharyngioma predicted to be BRAF V600E mutant (version 04, September 20, 2025) and eCRF (version V1.0, October 22, 2025). This statistical analysis plan will be revised in accordance with the revision of the study protocol.

The statistical analysis plan must be finalized after obtaining the sponsor's approval before the final data is locked.

### 2 Experimental purpose

This study aims to determine BRAF V600E mutation status in patients with newly diagnosed craniopharyngioma through an integrated predictive model of cerebrospinal fluid genomics and radiomics before surgery. For patients with papillary craniopharyngioma identified as having BRAF V600E mutation before surgery, dabrafenib + trametinib is used before surgery followed by elective surgery. The BRAF V600E mutation genotype in tumor tissue, total resection rate, complication rate, long-term recurrence rate, quality of life and other indicators after surgery were compared to evaluate the effectiveness of the preoperative integrated prediction model, the feasibility and necessity of preoperative use of targeted drugs, and the choice of surgical timing after medication.

### 3 Trial Design

#### 3.1 Trial Design

This study utilizes imaging data and cerebrospinal fluid collected prior to enrollment to predict tumor subtype in patients with craniopharyngioma. Patients predicted to have papillary craniopharyngioma receive investigational drug therapy in

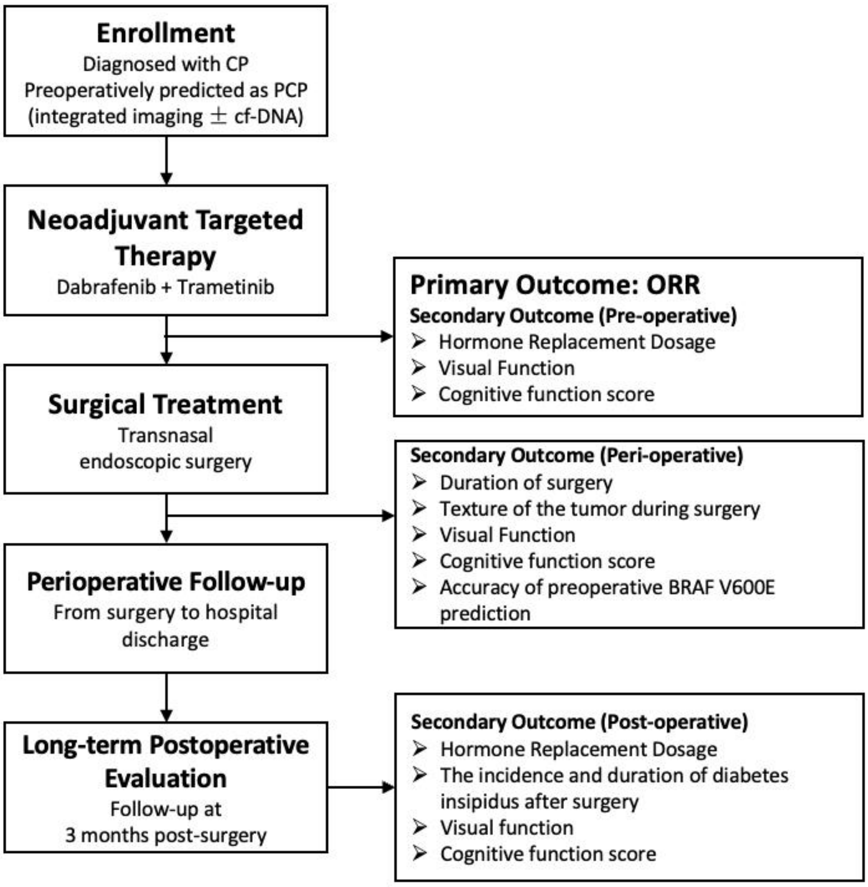

a single-arm, open-label design.

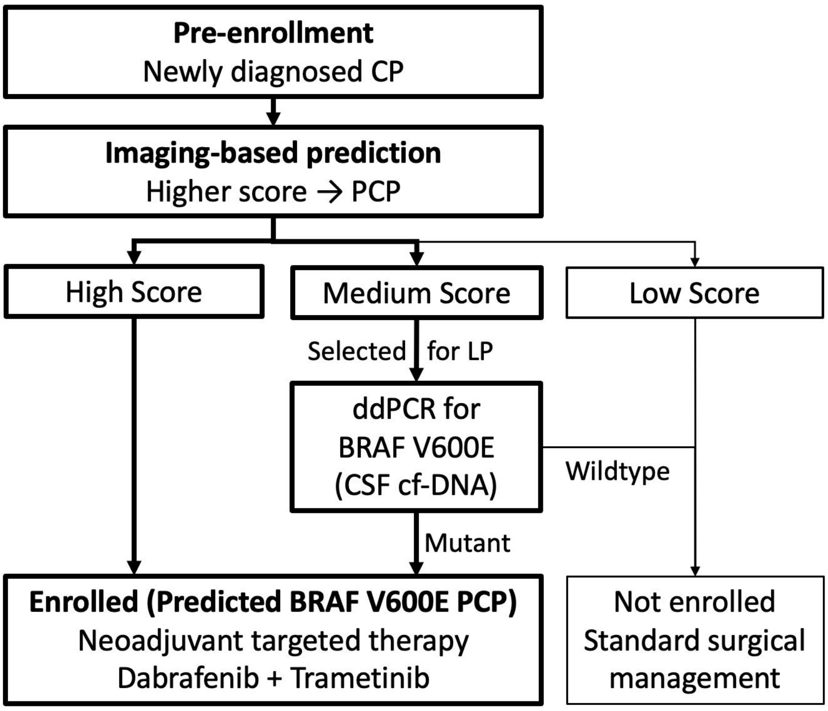

**Figure 1 Technical roadmap**

**Test process chart**

| Research Progress | Screening period | Treatment period | | | | | Surgery | Withdrawal | 3-month postoperative follow-up |
|---|---|---|---|---|---|---|---|---|---|
| | D-28~D-1 | D7 (±4) | D14 (±5) | D28 (±12) | D56 (±14) | D84 (±28) | | | 90 days (±45 days) after surgery |
| Informed consent | X | | | | | | | | |
| Demographic data | X | | | | | | | | |
| Height weight | X | ☐ | ☐ | ☐ | ☐ | ☐ | | | X |
| Craniopharyngioma diagnosis | X | | | | | | | | |
| History of radiotherapy for craniopharyngioma | X | | | | | | | | |
| Surgical history related to craniopharyngioma | X | | | | | | | | |
| History of craniopharyngioma | X | | | | | | | | |
| History of non-study tumors | X | | | | | | | | |
| History of smoking | X | | | | | | | | |
| History of alcohol consumption | X | | | | | | | | |
| Other Personal Histories | X | | | | | | | | |
| Physical examination | X | X | X | X | X | X | | X | X |
| Vital signs | X | X | X | X | X | X | | X | X |

| **Research Progress** | **Screening period** | **Treatment period** | | | | | **Surgery** | **Withdrawal** | 3-month postoperative follow-up |
|---|---|---|---|---|---|---|---|---|---|
| | **D-28~D-1** | **D7 (±4)** | **D14 (±5)** | **D28 (±12)** | **D56 (±14)** | **D84 (±28)** | | | **90 days (±45 days) after surgery** |
| 24-hour urine volume | X | | | | | | | X | X |
| Blood routine | X | X | X | X | X | X | | X | X |
| Urine routine | X | | | | | | | | X |
| Blood biochemistry | X | X | X | X | X | X | | X | X |
| Coagulation function | X | X | X | X | X | X | | X | X |
| Pituitary and related endocrine hormones | X | X | X | X | X | X | | X | X |
| Serum pregnancy test | X | | | | | | | | |
| Hormone replacement dose | X | X | X | X | X | X | | X | X |
| NYHA Cardiac function classification | X | X | X | X | X | X | | X | X |
| 12-lead electrocardiogram | X | | | | | | | X | X |
| Ophthalmic visual field examination | X | | | | | | | X | X |
| Imaging tests | X | X | X | X | X | X | | X | X |
| KPS score | X | | | | | | | X | X |
| Psychological assessment (MMSE Scale) | X | | | | | | | X | X |
| Entry and discharge | X | | | | | | | | |

| Research Progress | Screening period | Treatment period | | | | | Surgery | Withdrawal | 3-month postoperative follow-up |
|---|---|---|---|---|---|---|---|---|---|
| | D-28~D-1 | D7 (±4) | D14 (±5) | D28 (±12) | D56 (±14) | D84 (±28) | | | 90 days (±45 days) after surgery |
| standards | | | | | | | | | |
| Screening Conclusions | X | | | | | | | | |
| Surgical Records | | | | | | | X | | |
| Administration Record | | X | X | X | X | X | | | |
| Toxicity evaluation | | X | X | X | X | X | | | |
| Adverse events | | X | X | X | X | X | | X | X |
| Death information | | | | | | | | | X |
| Postoperative follow-up | | | | | | | | | X |
| Previous/combined medication | X | X | X | X | X | X | | X | X |
| Previous/combined non-pharmacological treatments | X | X | X | X | X | X | | X | X |
| The study ended | | | | | | | | X | |

Notes:

1) The items marked with "X" in this flowchart are the ones that must be improved, and the items marked with "☐" should be evaluated in combination with the clinical manifestations, economic conditions and wishes of the subjects;

2) Demographic information includes: date of birth, age, gender, ethnicity;

3) Vital signs include: body temperature; Respiratory rate; Pulse; Systolic blood pressure (sitting position); Diastolic blood pressure (sitting);

4) Physical examination should focus on the subject: skin and mucous membranes, lymph nodes, head, neck, chest, abdomen, spine, limbs, nervous system;

5) The blood routine includes: White blood cell count, red blood cell count, hemoglobin, platelet count, mean red blood cell hemoglobin concentration, mean red blood cell volume, red blood cell distribution width, red blood cell distribution width - coefficient of variation, platelet distribution width, mean platelet volume, large platelet ratio, neutrophil %, lymphocyte %, monocyte %, eosinophl %, basophl %, absolute neutrophil count, absolute lymphocyte count, absolute monocyte count, absolute eosinophil count, absolute basophhil count, neutrophil ratio, platelet-lymphoid ratio;

6) Urine routine includes: turbidity, color, urobilinogen, occult blood, bilirubin, ketone bodies, glucose, protein, pH, nitrite, leukocyte lipase, urine specific gravity, epithelial cells, red blood cell count, white blood cell count, pathological casts, small round epithelial cells, yeast, crystallization examination, bacteria, hyaluronic casts;

7) Coagulation function includes: prothrombin time, international normalized ratio, partial thromboplastin time, thrombin time, fibrinogen quantification, D-dimer, fibrinogen degradation products;

8) Blood biochemistry includes: alanine aminotransferase, aspartate aminotransferase, total protein, albumin, total bilirubin, direct bilirubin, cholinesterase, urea, creatinine, uric acid, potassium, sodium, chloride, blood calcium, inorganic phosphorus, blood magnesium, and carbon dioxide binding capacity.

9) Pituitary gland and related endocrine hormones include: adrenocorticotropic hormone, cortisol, thyroid-stimulating hormone, free thyroxine, free triiodothyronine, thyroid hormone, triiodothyronine, progesterone, estradiol, dehydroepiandrosterone sulfate, testosterone, luteinizing hormone, follicle-stimulating hormone, human growth hormone, insulin-like growth factor 1, prolactin;

10) Serum pregnancy test is only for women of childbearing age and is used to exclude patients who are pregnant;

11) Hormone replacement dose refers to the record of the dose and frequency of hormone replacement drugs such as glucocorticoids, thyroid hormones, and desmopressin used by the subjects during the evaluation period to replace their own reduced pituitary function;

12) Ophthalmic visual field examination includes binocular BCVA and binocular VFI, MD30-2, PSD30-2;

13) Imaging examination should be evaluated: target lesion location and target lesion size, target lesion size calculated according to two-dimensional measurement of tumor lesion (two-dimensional measurement of tumor lesion:) Take the maximum cross-section of the target lesion, measure the longest diameter line of the target lesion as the maximum diameter, and measure the longest diameter line of the target lesion in the direction perpendicular to the maximum diameter as the vertical diameter, expressing the size of the target lesion in the form of "maximum diameter × vertical diameter").

#### 3.2 Control type

This study will be designed with one arm and no control will be set.

#### 3.3 Investigational drugs

**Investigational Drug 1**

Generic Name Dabrafenib Tablets (BRAF Inhibitor)

Trade name: Tafinlar

English name: Dabrafenib

Properties: This product is a capsule and the contents are white powder

Specification: 75mg×120 capsules

Made by GlaxoSmithKline Manufacturing S.p.A

Registration numbers for imported drugs: H20190066, H20190067

**Investigational Drug II**

Generic Name Trametinib Tablets (MEK Inhibitor)

Trade name: Mekinist

English name Trametinib

Characteristics This product is a double-sided convex film coated tablet

Specification: 2mg×30 tablets

Produced by GlaxoSmithKline Manufacturing S.p.A

Registration numbers for imported drugs: H20190068, H20190069

#### 3.4 Dosing regimen

Treatment period: Patients receive dabrafenib 150mg po bid, trametinib 2mg po qd, with a 28-day treatment cycle for a total of 3 courses. Follow-up will be conducted on day 7/ 14/ 28/ 56/ 84 of the drug treatment until the patient underwent surgical treatment and the medication was discontinued.

Surgical period: During the visit, subjects who meet the preconditions for surgical treatment will receive surgical treatment at our center. Intraoperative tumor samples and surgery-related information of the subjects will be collected during the operation.

Follow-up period (3 months after surgery) : Subjects who underwent surgery after completing preoperative targeted adjuvant therapy should be followed up 3 months after surgery.

#### 3.5 Randomization method

This study will be a single-arm design and no randomization will be performed.

#### 3.6 Blinded

This study is a single-arm open-label trial and blinding is not applicable.

#### 3.7 Basis for sample size determination

This study is a single-arm exploratory study of the predicted BRAF V600E for the new onset of papillary craniopharyngioma. The sample size of this study was determined primarily based on the rarity of the disease, previous research data, and the feasibility of the study, rather than on event-driven statistical assumptions. Study completion is not contingent upon the occurrence of a prespecified number of progression events. The primary endpoint is objective response rate (ORR). Previous studies suggested an objective response rate of 80% for dabrafenib combined with trametinib.

Based on this, the Simon two-stage design is used in this study, with Class I error set at 0.05 and statistical power set at 0.8, assuming an objective response rate of less than 50% when the drug is ineffective and greater than 80% when the drug is effective. According to the PASS (V21 version) software, the minimax design is used: in the first stage, the drug is administered to 6 subjects, and the study is terminated if the number of cases reaching the endpoint event is less than 3. If the trial progresses to Phase 2, a total of 12 subjects will be required for both phases.

Given the low incidence of craniopharyngioma and based on our experience, we plan to subsequently include 40 patients with craniopharyngioma, use the previous model for prediction, and conduct experimental drug treatment for those predicted to have papillary type. Based on clinical practice, papillary craniopharyngioma patients account for approximately 30% of all craniopharyngioma patients. However, since a small number of papillary craniopharyngiomas share certain similarities in imaging features with adamantinomatous craniopharyngioma, in order to better study the effect of BRAF mutation inhibitors as adjuvant treatment for papillary craniopharyngioma, it is expected that about 12 patients will be included in the drug treatment It will meet the requirements of the initial exploratory study.

### 4 Evaluation metrics

#### 4.1 Demographic and baseline characteristics

- Demographic indicators: Age (in years) ((informed consent date - date of birth +1) /365.25), gender, ethnicity, height, weight, etc.
- Diagnosis and treatment of craniopharyngioma: Diagnosis of craniopharyngioma, disease duration ((date of enrollment - date of first diagnosis +1) /30.4375, unit: months), history of radiotherapy, related surgical history, etc.
- Previous medical history: Other medical history, smoking history, drinking history, other personal history, etc.
- Treatment history: Previous medication use, previous non-pharmacological treatment, etc.
- Screening criteria: Serum pregnancy test, cerebrospinal fluid collection, etc.

#### 4.2 Effectiveness estimation targets

**The validity evaluation criteria:** Tumor response will be assessed according to RANO criteria in this study. The study was a single-center study, and all imaging assessments were independently interpreted by two imaging experts with extensive experience in diagnosing and treating skull base tumors, with blinding of clinical information and treatment outcomes throughout the interpretation process. If there is a significant difference in the determination of tumor volume change or remission grade between the two imaging assessors, a third imaging expert will review and make the final decision. All imaging assessments were conducted in accordance with the uniform assessment criteria pre-set in the study protocol to ensure consistency and reliability of the assessment results.

| | Complete remission (CR) | Partial remission (PR) | Stable disease (SD) | Progress disease (PD) |
|---|---|---|---|---|
| T1+ enhancement | Not seen | Reduction ≥50% | -50% to +25% | Increase ≥25% |
| New lesions | Not seen | Not seen | Not seen | Visible |
| clinical picture | Stable/Improved | Stable/Improved | Stable/Improved | Deterioration* |
| Judgment conditions | All above | All above | All above | All above |

RANO-based Response Criteria

*KPS score reduced from 100 or 90 to ≤70, reduced by more than 20 points from 80 or 70; Or there is a significant decline in visual field.

###### 4.2.1 Validity evaluation time

Tumor imaging tests will be conducted during the screening period, D7, D14, D28, D56, D84, early withdrawal/exit from the group, and postoperative follow-up.

##### 4.2.2 Primary estimated targets

**Objective response rate (ORR):** ORR is defined as the proportion of patients achieving complete remission (CR) or partial remission (PR) at the prespecified primary response assessment, defined as the MRI assessment at the end of neoadjuvant therapy (D84). For participants who undergo surgery earlier than D84, the last MRI assessment prior to surgery will be used as the primary response assessment.

##### 4.2.3 Secondary estimated targets

- **Progression-free survival**: The time from the first study of drug treatment to PD or death (whichever occurs first). Surgery will not be considered a progression event. Patients without documented progression or death will be censored at the date of last disease assessment within the preoperative observation window.

- **Local control rate**: The proportion of subjects who achieved complete response (CR), partial response (PR), or stable disease (SD) after treatment.
- **HPA and HPT axis replacement therapy doses:** Compare the pituitary-adrenal and thyroid replacement therapy doses (dose and frequency of cortisone and levothyroxine) in subjects before treatment, before surgery, and 3 months after surgery.
- **Visual function:** Compare visual field changes in subjects before treatment, before surgery, and 3 months after surgery.
- **Cognitive function score:** Compare the changes in MMSE scores of the subjects before treatment, before surgery, and 3 months after treatment.
- **Accuracy of preoperative prediction of BRAF** mutations: Comparing the consistency of BRAF V600E mutants in postoperative tumor pathology reports with preoperative multimodal predictions.
- **Incidence and duration of diabetes insipidus after surgery:** Fluid intake and output, changes in blood sodium, dose of desmopressin replacement therapy for 24 hours one week after surgery in the subjects.
- **Duration of the operation:** The duration of the operation in the subjects.

#### 4.3 Exploratory indicators

- **Intraoperative tumor blood supply texture:** Tumor texture in subjects during the operation.
- **Post-treatment tumor epigenetics:** Epigenetic profiles of tumor specimens obtained by subjects during the operation.

#### 4.4 Safety evaluation indicators

- Adverse events;
- Laboratory tests: Baseline, D7, D14, D28, D56, D84, early withdrawal/exit from the group, and postoperative follow-up with laboratory tests such as blood routine, urine routine, and blood biochemistry;

**Blood routine:** White blood cell count, red blood cell count, hemoglobin, platelet count, mean red blood cell hemoglobin concentration, mean red blood cell volume, red blood cell distribution width, red blood cell distribution width - coefficient of variation, platelet distribution width, mean platelet volume, large platelet ratio, neutrophil ratio, lymphocyte ratio, monocyte ratio, eosinophl ratio, basophl ratio, absolute neutrophil count, absolute lymphocyte count, absolute monocyte count, absolute eosinophil count, absolute basophhil count, neutrophil ratio, platelet-lymphoid ratio;

**Urine routine:** turbidity, color, urobilinogen, occult blood, bilirubin, ketone bodies, glucose, protein, pH, nitrite, leukocyte lipase, urine specific gravity, epithelial cells, red blood cell count, white blood cell count, pathological casts, small round epithelial cells, yeast, crystallization examination, bacteria, hyaluronic casts;

**Blood biochemistry:** Alanine aminotransferase, aspartate aminotransferase, total protein, albumin, total bilirubin, direct bilirubin, cholinesterase, urea, creatinine, uric acid, potassium, sodium, chloride, blood calcium, inorganic phosphorus, blood magnesium, carbon dioxide binding capacity;

**Coagulation function:** Prothrombin time, international normalized ratio, partial thromboplastin time, thrombin time, fibrinogen quantification, D-dimer, fibrinogen degradation products;

**Pituitary gland and related endocrine hormones:** adrenocorticotropic hormone, cortisol, thyroid-stimulating hormone, free thyroxine, free triiodothyronine, thyroid hormone, triiodothyronine, progesterone, estradiol, dehydroepiandrosterone sulfate, testosterone, luteinizing hormone, follicle-stimulating hormone, human growth hormone, insulin-like growth factor 1, prolactin;

- Vital signs: Baseline, D7, D14, D28, D56, D84, early withdrawal/exit from the group, postoperative follow-up for vital sign examination of body temperature, pulse, respiratory rate, systolic blood pressure, diastolic blood pressure, etc.
- Physical examination: Physical examination of skin and mucous membranes, lymph nodes, head, neck, chest, abdomen, spine, limbs, nervous system, etc. at baseline, D7, D14, D28, D56, D84, early withdrawal/exit from group, postoperative follow-up;
- 12-lead electrocardiogram examination: QT, heart rate, QTc, etc. electrocardiogram examination at baseline, early withdrawal/exit from group, and postoperative follow-up. Visits such as D7, D14, D28, D56, and D84 are determined based on the actual situation.

### 5 Statistical analysis of the population

Before the database is locked, the subjects for each statistical analysis population are determined through joint discussion by the sponsor and the statistician at the data review meeting.

Full analysis set (FAS) : including all enrolled subjects. The FAS will serve as the primary analysis population for efficacy analyses and will also be used for the analysis of baseline characteristics.

Safety data set (SS) : Including all subjects who are enrolled, received at least one trial drug, and had at least one post-administration safety evaluation, the safety dataset will be used for the safety analysis.

Evaluable Set (EVS) : Includes all participants who are predicted to have the BRAF V600E genotype and had received at least one study drug, had baseline measurable lesions and had at least one post-treatment tumor assessment.

Per-protocol set (PPS): The per-protocol set is a subset of the FAS, including all enrolled subjects with no protocol deviations that significantly affected the primary efficacy assessment. Analyses based on the PPS will be performed as supportive sensitivity analyses.

### 6 Phase analysis

The phase determination of continuation/termination will be made after the completion of the first phase in accordance with the Simon two-phase design preset in the scheme.

### 7 Quality assurance of data transfer, computation, and reporting processes

After the database is locked, the eCRF tabulated data will be exported by the data management team from the EDC system and sent to the statistical programming team in the form of SAS datasets. Medical coding and protocol deviation data will be sent by the data management team to the statistical programming team in EXCEL or Word format. The results of all statistical analyses were verified through two-person independent programming, and the results were consistent. The test process and anomalies in the statistical analysis will be reflected in the statistical analysis report, and the statistician will review and confirm the content of the statistical analysis report.

### 8 Data processing principles

#### 8.1 Baseline

A baseline is defined as the last non-missing observational data (including unplanned checks) before the first administration.

#### 8.2 Study Day

Study day: The number of days for efficacy assessment/safety assessment relative to the reference date. The date of first administration serves as the reference date. The reference date day will be counted as Day 1.

The study day is calculated as follows:

If the assessment/examination is before the reference date, then "study date = assessment/examination date - reference date".

If the assessment/examination is after or on the reference date, then "Study date = assessment/examination date - reference date +1".

#### 8.3 Missing value handling

##### 8.3.1 AE Date Missing

- When analyzing AE, it is necessary to compare the start date of the adverse event with the first administration date. If the start/end date of the adverse event is missing or partially missing, fill in the following principles:

| Date | Known (missing) | Filling rules |
|---|---|---|
| Start Date | NA (absent YMD) | Date of first administration |
| | YYYY (MD missing) | If YYYY< YYYY of the first administration date, fill in YYYY-12-31 |
| | | If YYYY= YYYY of the date of first administration, fill in the date of first administration |
| | | If YYYY> YYYY of the date of first administration, fill in YYYY-01-01 |
| | YYYY-MM (missing D) | If YYYY-MM< YYYY-MM on the first administration date, fill it with YYYY-MM-DD (D is the last day of M month). |

- If the padded start date is after the end date, the end date is taken as the corresponding start date.
- If the filled end date is after the last visit date, the last visit date shall be the corresponding end date, and if it is after the date of death, the date of death shall be the corresponding end date.
- List according to the original data.

##### 8.3.2 Missing dates of previous/concomitant medications and previous/concomitant non-pharmacological treatments

- When analyzing concomitant/non-pharmacological treatments, if the start/end dates of medication use are missing or incomplete, fill them in according to the following logic.

| Date | Missing | Filling rules |
|---|---|---|
| Start Date | NA (absence of YMD | No filling, all should be recorded as previous medication/non-medication treatment |
| | YYYY (MD missing) | YYYY-01-01 |
| | YYYY-MM (missing D) | YYYY-MM-01 |
| End Date | NA (absent YMD) | Not filled, all should be recorded as combined medication/non-drug treatment |
| | YYYY (MD missing) | YYYY-12-31 |
| | YYYY-MM (lacking D) | YYYY-MM-DD (D is the last day of M month) |

- If the end date after filling is still not determined to be before the first administration date, it will be considered a combination/non-pharmacological treatment.
- If the end date after filling is after the date of the last visit, the date of the last visit is taken as the corresponding end date; if it is after the date of death, the date of death is taken as the corresponding end date.
- List according to the original data.

##### 8.3.3 The date of the disease course is missing

When calculating the course of craniopharyngioma, if there is a missing date for the first diagnosis of the craniopharyngioma, the following rules will be used to fill in the missing date, and if the filled date is later than the enrollment date, the enrollment date will be used. The imputation rules are as follows:

- Years, months, and days are missing and not filled in;
- Missing months and days: a) If the first diagnosis date is the same as the enrollment date year (YYYY), fill in the middle date between January 1 of the first diagnosis date year (YYYY-01-01) and the enrollment date, and if the middle date is not an integer date but between two dates, fill in the first date; b) Fill in July 1 (YYYY-07-01) if the first diagnosis date is different from the enrollment date year;

- Missing days: a) If the date of first diagnosis is the same as the year and month (YYYY-MM) of the enrollment date, fill in the middle date between the 1st day of the month of the first diagnosis (YYYY-MM-01) and the enrollment date, and fill in the first date if the middle date is not an integer date but between two dates; b) If the date of first diagnosis is different from the year and month of the enrollment date, fill in the 15th day of the month (YYYY-MM-15).
- List based on the original data.

For the remaining demographic, baseline, safety and efficacy indicators, unless otherwise specified, all missing data should not be filled in in any way. Statistical description should be treated as missing data, and listing should be described as EDC content.

#### 8.4 Processing of unused data, illogical data, etc

All data will be used for statistical analysis or listing, unless otherwise specified.

Illogical data will be discussed and decided at the data review meeting whether to exclude or conduct sensitivity analysis on a case-specific basis.

### 9 Statistical analysis methods

#### 9.1 General Principles

Statistical analyses will be performed using SAS software (version 9.4 or above).

Descriptive analysis will be based on the following principles: continuous variables will be described using the number of non-missing observations, mean, standard deviation, median, interquartile range (Q1, Q3), minimum, and maximum; Categorical variables will be statistically described using the number of cases in each category and their percentages.

The data used in general statistical tables will be presented in a list; Unless otherwise specified, all lists will be sorted by filter number, check item, visit, etc.

Unless otherwise specified, all statistical tests use two-sided tests, and a p value less than 0.05 is considered statistically significant.

During the calculation process, the original data is used for all calculations. Only when the final data is presented, rounding is adopted. The retention rules for decimal places are as follows: No more than five decimal places will be retained.

| Statistics | Retain the number of decimal places |
|---|---|
| Minimum, maximum | Be consistent with the original record, which includes eCRF and external data results |
| Mean, standard deviation, median, coefficient of variation, and confidence interval | Keep two decimal places |
| Q1, Q3 | 2 bits more than the original recorded data |
| Percentages and their confidence intervals (%) | Keep one decimal place; If the frequency is 0, do not report the percentage; if the percentage is 100%, report the percentage as 100, do not retain decimal places |
| P value | Keep four decimal places for values greater than or equal to 0.0001; P values less than 0.0001 are indicated as "<.0001"; When the P value is greater than 0.9999, use ">0.9999"; If the P value is close to 0.05 (rounded to 0.05), show the cutoff value without rounding, for example: 0.049999, show 0.0499. |
| Statistics t values, F values, etc | Keep four decimal places |

#### 9.2 Distribution of subjects

- Use tables and flowcharts to describe the screening, enrollment, administration situations and the number and percentage of subjects who completed the study.
- Use tabular grouping to describe the number and percentage of subjects in each analysis dataset;
- Provide a list of subjects whose screening failed, a list of subjects who dropped out of the trial and the reasons for their withdrawal, and a list of subjects who were not included in the analysis set.
- A summary analysis and checklist of protocol deviations.
  - Surgery was allowed to be performed early when the subjects achieved the preset tumor response criteria during the study drug treatment and the investigator determined that they were suitable to enter the surgical stage. Such early surgery is considered a treatment path that conforms to the study

protocol and is not treated as a protocol deviation.

#### 9.3 Analysis of demographic data and baseline characteristics

- FAS analysis will be used;
- Statistically describe demographic data and other baseline characteristics.

#### 9.4 Analysis of medication adherence and concomitant medication

##### 9.4.1 Medication compliance

- FAS analysis will be used.
- Aggregate actual drug exposure, dosing compliance, etc.

  Medication compliance (%) = (actual total dosage mg/ planned total dosage mg) *100%. The actual total dosage is the information collected from the database. The planned total dosage of dabrafenib =150mg*2* duration of administration. The planned total dosage of trametinib =2mg* duration of administration. Duration of administration = date of last administration - date of first administration +1. And summarize the number and percentage of subjects with medication compliance < 80%, 80% - 120%, > 120%.
- Summarize the number and percentage of participants with dose adjustments, permanent discontinuation, and suspension of administration.
- Summarize the actual number of days of medication use, the actual administration intensity and the relative administration intensity, etc. The actual number of days of dabrafenib administration (days) = actual number of administrations /2; Trametinib actual days of administration (days) = actual number of administrations. Actual dosage intensity (mg/ day) = actual total dosage (mg)/duration of administration; Planned administration intensity (mg/ day) = planned daily dosage (dabrafenib: 150mg*2, trametinib: 2mg); Relative administration intensity (%) = actual administration intensity (mg/ day)/planned administration intensity (mg/ day) *100%.

##### 9.4.2 Combination/non-drug treatment analysis

Combination therapy includes previous medications and concomitant medications.

Previous medication refers to non-study medication that will be initiated and ended before the first use of the investigational drug.

The study only allowed concomitant administration for the management of adverse events or the treatment of sudden illness. Concomitant medication is a non-study medication that meets one of the following conditions:

- Initiated after the first use of the investigational drug
- Start before the first use of the investigational drug and continue to use it after the first use.

The names of concomitant medications are encoded using WHOCC (ATC/DDD Index 2025 or above) and the anatomical - Therapeutic - Chemical drug Classification System (ATC), and summarized by drug classification (ATC1 and ATC4).

A checklist will be provided for all comorbidities/non-drug treatments.

#### 9.5 Efficacy estimation target analysis

##### 9.5.1 Primary estimation objective analysis

- **Testing hypotheses:** This study is an exploratory trial and no statistical inferences are made.
- **Main estimation method**

The tumor objective response rate (ORR) at the prespecified primary response assessment (Day 84 or last MRI prior to surgery), including the proportion of complete response (CR) to partial response (PR) (CR+PR). Patients who did not undergo any assessment will be considered non-responders. After each assessment, the follow-up regimen follows the following approach:

- If rated as PD, withdraw from the study and undergo surgery immediately.
- If the patient's tumor significantly shrinks by more than 50% during the treatment period (i.e. CR or PR), withdraw from the study and undergo surgery immediately.
- If the tumor maintains SD during the medication period, the patient will receive surgical treatment as planned after one month of medication ends.

Based on FAS, PPS, and EVS, the number and percentage of subjects with objective response were aggregated, and the 95% confidence interval of the objective response rate will be calculated using the Clopper-Pearson method.

Swimlane diagrams will be drawn for the overall tumor evaluation results of different subjects at each visit point.

##### 9.5.2 Secondary estimation target analysis

1） Progression-free survival: Based on FAS and EVS, the time from the first study drug treatment to PD or death (whichever occurs first). Surgery marks the end of preoperative observation and will not be treated as a censoring event. Given that the investigational drug treatment in this study design will uniformly enter the surgical stage and the expected rate of tumor progression events is extremely low, the progression-free survival (PFS) analysis was mainly used as a descriptive exploratory analysis. Kaplan-Meier estimates and median PFS will be reported only if estimable. In such cases, PFS will be reported using event counts and the distribution of observed follow-up time (time at risk) from treatment initiation to progression/death, surgery, or last assessment, as applicable. The date of surgery will be treated as the end of the preoperative observation period rather than a censoring event for survival estimation. The event occurrence and the distribution of follow-up time (months) = (withdrawal/surgical start date - first study drug treatment date +1) /30.4375.

2) Local control rate: The proportion of subjects whose overall response was CR, PR, or SD (i.e. CR+PR+SD). Based on FAS and EVS, summarize the number and percentage of subjects with local control and their 95% confidence intervals.

3) HPA and HPT axis replacement therapy doses: Compare the doses of pituitary-adrenal and thyroid replacement therapy (dose and frequency of cortisone and promethole) in subjects before treatment, before surgery, and 3 months after surgery. Based on FAS, hormone replacement therapy doses were described by visit. Paired t-tests/sign-rank sum tests were used for quantitative data to compare changes from baseline after treatment, and a line graph of mean changes over time was plotted.

4) Visual function: Compare the changes in visual field of the subjects before treatment, before surgery, and 3 months after treatment. Based on FAS, by visiting descriptive visual acuity and visual field, etc., paired t-test/sign-rank sum test was used to compare changes from baseline after treatment, and a line graph of mean over

time was plotted; The clinical determination of the examination results was described in the form of a cross-table before and after.

5) Cognitive function scores: Compare the changes in MMSE scores of the subjects before treatment, before surgery, and 3 months after treatment. Based on FAS, descriptive MMSE scores by visit were compared from baseline after treatment using paired t-test/signal-rank sum test, and a line graph of mean over time was plotted.

6) Accuracy of preoperative prediction of BRAF mutations: Based on FAS, the Kappa coefficient was used to assess the consistency of postoperative tumor pathology reports of BRAF V600E mutants with preoperative multimodal predictions in subjects.

7) Incidence and duration of diabetes insipidus after surgery: Based on FAS, summarize the 24-hour fluid intake and output, changes in blood sodium, and dose of desmopressin replacement therapy in subjects for one consecutive week after surgery.

8) Duration of surgery: Based on FAS, summarize the duration of surgery for the subjects; Operation duration (h) = operation end date time - operation start date time, unit: h, rounded to two decimal places.

##### 9.5.3 Exploratory estimation target analysis

1) Intraoperative tumor blood supply texture: Based on EVS, summarize the number and percentage of cases of intraoperative tumor texture in the subjects.

2) Post-treatment tumor epigenetics: Based on EVS, summarize the number and percentage of cases of epigenetics of tumor specimens obtained during surgery in subjects.

#### 9.6 Safety analysis

- The security analysis will be based on the Safety data set (SS).

##### 9.6.1 Adverse events

All adverse events (AEs) will be classified by system organ classification (SOC) and preferred term (PT) in accordance with the Dictionary of Regulated Activity Medicine (MedDRA 28.1 or above).

Adverse events that occur during treatment are defined as those that occur or worsen after the first administration of the investigational drug. The occurrence of adverse events requires the investigator to distinguish: related to drug treatment/related to surgical treatment. In this clinical trial, only the adverse events that occurred during the treatment period were statistically described. Adverse events that occurred before the first administration are only shown in the list of adverse events.

An adverse event related to the study drug is defined as an adverse event whose relationship with any study drug is determined to be "definitely related", "possibly related", or "undetermined".

A serious adverse event (SAE) is defined as an adverse medical event at any dose that leads to death; Life-threatening (referring to the immediate risk of death in a serious patient, not the assumption that death may occur in the future if it develops seriously); Need for hospitalization or an extension of existing hospital stay; Permanent or significant loss of function; Congenital malformations or birth defects; It leads to other significant medical events that, if left untreated, may occur as listed above. "Whether it is a serious adverse event" select "Yes" for adverse events.

- Summarize the number of cases, and incidence rates of the following adverse events: Adverse events, adverse events related to the study drug, adverse events related to dabrafenib, adverse events related to trametinib, adverse events related to surgical treatment, serious adverse events, serious adverse events related to the study drug, serious adverse events related to surgical treatment, adverse events leading to withdrawal, adverse events related to the study drug leading to withdrawal Events, adverse events related to surgical treatment that led to withdrawal, adverse events of severity ≥3, adverse events of severity ≥3 related to the study drug, adverse events of severity ≥3 related to surgical treatment, toxic reactions, toxic reactions related to the study drug.
- The number of cases, and incidence rates of the following adverse events were aggregated by SOC and PT: Adverse events, adverse events related to the study drug, adverse events related to surgical treatment, serious adverse events, serious adverse events related to the study drug, serious adverse events related to surgical treatment, adverse events leading to withdrawal, adverse events related to the study drug leading to withdrawal, adverse events related to surgical

treatment leading to withdrawal, severity ≥3 Grade adverse events, grade ≥3 adverse events related to the study drug, grade ≥3 adverse events related to surgical treatment, toxic reactions, toxic reactions related to the study drug;

- The number of cases, the number of cases, and the incidence of the following adverse events by severity, SOC, and PT: Adverse events, drug-related adverse events, surgery-related adverse events.
- A detailed list of all adverse events, adverse events related to the study drug, adverse events related to surgical treatment, serious adverse events, and toxic reactions.

##### 9.6.2 Laboratory tests

- Describe the clinical determination of laboratory test results using cross-tabular forms before and after;
- Summarize the changes from baseline to the most severe results after baseline using the cross-tabular method (take the most severe results in the order of abnormal with clinical significance > abnormal without clinical significance > normal > unexamined). The most severe results in the cross table will be included in the planned field visit;
- The diachronic variation and relative baseline variation of quantitative results from laboratory tests;
- List the subjects' examination items list of abnormal items before and after treatment;
- List the previous clinically significant changes in the subjects' test item abnormalities;
- A detailed list of all the laboratory test results of the subjects.

##### 9.6.3 12-lead electrocardiogram examination

- Describe the results of the 12-lead electrocardiogram using a cross-tabulation of the front and back;
- The changes in the most severe results after baseline in the 12-lead electrocardiogram results were summarized using the cross-tabulation method (the most severe results were selected in the order of abnormal with clinical significance > abnormal without clinical significance > normal > unexamined). The most severe results in the cross table will be included in the planned visit;

- List the items that are abnormal before and after treatment for the subjects;
- List the previous changes in the subjects' examination items that were clinically significant;
- The diachronic and relative baseline changes in quantitative results of 12-lead electrocardiograms;
- A detailed list of 12-lead electrocardiogram results for all subjects.

##### 9.6.4 Physical examination

- Describe the results of the physical examination using cross-tabulation tables before and after;
- Summarize the changes from baseline to the most severe results after baseline using the cross-tabulation method (take the most severe results in the order of abnormal with clinical significance > abnormal without clinical significance > normal > unexamined). The most serious results in the cross table will be included in the planned visit;
- List the items that are abnormal after treatment for the subjects;
- List the previous changes in the subjects' examination items that were clinically significant;
- A detailed list of physical examination results for all subjects.

##### 9.6.5 Vital Signs

- The changes from baseline to the most severe results after the clinical determination baseline of the vital signs examination were summarized using the cross-table method before and after (the most severe results were selected in the order of abnormal with clinical significance > abnormal without clinical significance > normal > not checked). The most severe results in the cross table will be included in the planned visit;
- The diachronic variations and relative baseline changes of the quantitative results of measured values of various vital signs;
- List the previous changes in the subjects' examination items that were clinically significant;
- A detailed list of the results and anomalies of each vital sign examination.

##### 9.6.6 Other tests

- The clinical determination of NYHA cardiac function grading test results was described using a cross-tabular form before and after;

- Diachronic changes and relative baseline changes in weight test results;
- Summarizing postoperative follow-up, death information, and surgical complications of the subjects;
- Detailed list of NYHA cardiac function classification, height and weight, postoperative follow-up, death information, and surgical complications for all subjects.

### 10 Modified procedures for the original statistical analysis plan

Both clinical trial protocol and eCRF version updates may result in SAP updates, but SAP must be finalized until the database is locked. After the database is locked, all statistical analyses that deviate from the original statistical analysis plan will only be used as sensitivity analyses or supplementary analyses.
